# The Global Imbalance in Telemedicine Research: An Analysis of Knowledge Production and Socioeconomic Drivers

**DOI:** 10.64898/2026.02.27.26347284

**Authors:** Seyed Sahab Aarabi, Farbod Semnani, Mojtaba Sedaghat

## Abstract

**Background:** This study aims to explore disparities in telemedicine research, investigate the impact of the COVID-19 pandemic on these inequalities, and examine the association between various socioeconomic factors and telemedicine research output across Low- and Middle-Income Countries (LMIC) and High-Income Countries (HIC), and World Health Organization (WHO) regions.

**Methods:** A comprehensive search strategy was developed to identify telemedicine-related documents (2018–2022) in Scopus and SciVal, with false positives and negatives resolved. Mann-Whitney U and Wilcoxon Signed Rank tests compared publication volume and Field-Weighted Citation Impact (FWCI). A novel metric, Research Interest (RI), was calculated by dividing telemedicine publications by total outputs in medicine and life sciences. WHO regions were ranked using TOPSIS. Spearman Rank Correlation assessed links between socioeconomic variables and research output separately in HIC and LMIC. Analyses were conducted using R (v4.3.2).

**Results:** We retrieved 16,584 telemedicine-related articles: 4,244 from 58 LMIC and 13,622 from 47 HIC, including 1,282 collaborative publications (30% of LMIC and 9.4% of HIC outputs). HIC consistently produced more publications than LMIC. While FWCI differences were significant in the pre-COVID era (Cliff’s Delta = 0.48), no significant difference was observed post-COVID. RI for telemedicine showed no significant difference between HIC and LMIC in any timeframe. The Western Pacific led in quality metrics, while the Americas ranked highest overall. Southeast Asia ranked lowest in both. Exclusively among HIC, Health Expenditure (Purchasing Power Parity adjusted) (r = 0.63, r = 0.45) and Human Development Index (r = 0.50, r = 0.47) were moderately, and ICT service exports (USD) (r = 0.72, r = 0.33) were strongly correlated with both telemedicine scientific output and RI.

**Conclusion:** Global inequalities in telemedicine research favor HIC, though the gap narrowed post-COVID. Among HIC, telemedicine research patterns more proportionately reflect socioeconomic indicators, research capacity, infrastructure, and domestic health needs.

## 1. Introduction

The World Health Organization (WHO) defines telemedicine as delivering healthcare services over distance by medical experts. Information and Communication Technologies (ICT) play an important role in exchanging valid information for diagnosing, treating, and preventing diseases. The primary goal of telemedicine is to enhance the health and well-being of individuals and communities (1).

The use of telemedicine services has increased at an unprecedented rate in recent years, particularly with the emergence of the COVID-19 pandemic (2). The Centers for Disease Control and Prevention (CDC) reported that in 2021, 37% of American adults used telemedicine services, with higher adoption rates observed among older individuals, those with higher family incomes, greater educational attainment, and those living in urban areas (3). Additionally, by 2021, 52% of insurers worldwide had integrated telehealth services into their offerings (4).

Telemedicine allows cooperation between primary care physicians and specialists, reducing wait times for medical feedback, unnecessary transportation costs, and unrequired in-person referral examinations. In regions with limited access to specialized healthcare providers, it facilitates remote medical services and helps decrease overall wait times for care (5–10). Given these advantages, telemedicine plays a crucial role in global and public health.

Telemedicine is particularly important in Low- and Middle-Income Countries (LMIC), where a larger rural population, restricted access to primary and specialized healthcare services, a shortage of medical professionals, and the high cost of transportation pose significant barriers to healthcare access (11–15). Income level was reported as one of the factors affecting telemedicine services utilization (16). Despite its critical importance in LMIC, the adoption and utilization of telemedicine remain significantly lower than in High-Income Countries (HIC) (14). One contributing factor may be the inequality in telemedicine-related research between these countries (17–19). The role of knowledge production in exacerbating disparities within health systems is often neglected by researchers studying healthcare inequalities (20). Vulnerable populations not only have the least access to telemedicine services but also pay the least attention to conducting research tailored to their needs.

Several socioeconomic factors have been shown to influence scientific production across various fields (21–25). There are a few studies investigating the relationship between socioeconomic indicators and the production of telemedicine research. One study utilized the MEDLINE database to examine the correlation between telemedicine publications per million inhabitants (1964–2003) and factors such as population density, Gross National Product (GNP), Human Development Index (HDI), and the number of Primary Care Providers (PCP) per 1000 inhabitants (26). Another study reported that a higher HDI correlates with more teleophthalmology articles per million people (27).

Bibliometric analysis is a statistical method used to evaluate and quantify the growth and trends within a specific field of research (28). This method is particularly used to study topics experiencing exponential growth in publications. The field of telemedicine research has grown rapidly in recent years, notably with the emergence of the COVID-19 pandemic, demonstrating the necessity of conducting bibliometric studies in this field (29). A few bibliometric studies have studied the impact of COVID-19 on telemedicine publications (29–31).

To the best of our knowledge, this is the first study to comprehensively analyze the possible socioeconomic factors affecting knowledge production in telemedicine research, examining LMIC and HIC separately. Our findings aim to provide policymakers in each group with insights into these factors, helping to address existing disparities in telemedicine implementation and utilization. Besides, we represent the existing differences and inequalities in telemedicine research between HIC and LMIC, and across all WHO regions. Further, we assess the impact of the COVID-19 pandemic on telemedicine publications.

## 2. Materials and Methods

This research is an explorative study consisting of two main parts. In the first section, we descriptively and analytically compare chosen bibliometric indices of telemedicine-related documents between HIC and LMIC, pre-and post-COVID era, and across all WHO regions. In the second section, we performed a cross-sectional study to analyze the correlation between socioeconomic factors and bibliometric indices comparatively across LMIC and HIC.

### 2.1 Ethics Statement

This study was reviewed and approved by Research Ethics Committees of School of Medicine-Tehran University of Medical Sciences with the approval number: IR.TUMS.MEDICINE.REC.1401.673, dated 2023-01-16

### 2.2 Database selection

We used the Scopus database to retrieve telemedicine-related documents published between 2018 and 2022. We selected Scopus for its extensive publication coverage, including document-specific impact indicators through SciVal (Its companion online tool), and accessibility to researchers in LMIC through journals with lower impact factors (32,33). Its comprehensive scope across diverse disciplines makes it particularly suitable for analyzing interdisciplinary fields like telemedicine (34,35).

### 2.3 Search strategy

The search time frame was defined from 2018 to 2022 to uncover the role of the COVID-19 pandemic on telemedicine research and ensure an adequate number of articles retrieved for the relevant comparisons. Furthermore, a sufficient amount of time has elapsed since the publication of these articles, allowing for a more comprehensive assessment of their impact.

We developed the search string, whose rationale is available in the supplementary file. Keywords were obtained by reviewing MESH terms, telemedicine systematic reviews, and similar bibliometric studies (17,29,36,37). No language restrictions were applied, and only peer-reviewed studies were included. Hence, Conference papers, notes, editorials, erratums, letters, and book chapters were excluded. In addition, retracted documents were omitted from the study.

The search was performed and finalized on August 9, 2024. After applying the limitations, 16584 relevant articles were obtained from Scopus and exported as CSV files containing citation information and bibliographic data.

### 2.4 Search Validation

Considering the nature of bibliometric studies and the large number of articles retrieved, manually screening titles and abstracts was not feasible. As a result, we used a screening method proposed by the other bibliometric studies (36,38,39).

For false positive screening, two authors (S.S.A & F.S) reviewed the 100 most-cited articles regarding relevance to telemedicine. Initially, five irrelevant articles were found. However, after a proper modification of the search string, all articles were identified as relevant in the next review (see the final search string in the supplementary file).

For false negative (missing records) screening, the Scopus profiles of the 11 most prolific authors (those with the highest number of articles in our search) were reviewed to determine the number of telemedicine-related articles they had published from 2018 to 2022. A Spearman correlation test was then applied to assess the relationship between the number of articles found in our search and the Scopus profiles of the authors (Table S1). We discovered a significant positive correlation (r=0.736, P< 0.01), suggestive of the high validity of our search strategy.

### 2.5 Countries Classification

This study categorized the countries based on two important factors to achieve the stated objectives:

#### 2.5.1 Income level

The World Bank Group assigns the world’s economies into four income groups (low, lower-middle, upper-middle, and high) annually based on Gross National Income (GNI) per capita (40). In this study, low, lower-middle, and upper-middle-income countries were classified as LMIC and the rest as HIC. We retrieved countries’ income classes from 2018 to 2022 from the World Bank website and excluded countries lacking a complete income classification for these years. The final income category for each country was determined based on the most common income group they had been assigned to during this period.

#### 2.5.2 WHO Regions

We conducted another classification based on WHO regions for a more comprehensive comparison of telemedicine-related scientific production between countries (41). Furthermore, we carried out a separate analysis for Iran to evaluate the country’s performance in telemedicine research relative to other nations.

### 2.6 Timing

WHO declared COVID-19 a pandemic on March 11, 2020 (42). Thus, we split the study period into two parts (2018-2019 and 2020-2022) to observe the impact of the COVID-19 pandemic on telemedicine-related studies. The mentioned searches for LMIC and HIC were also retrieved separately for these two timeframes.

### 2.7 Bibliometric Indices

In Scopus and SciVal, countries are determined based on the authors’ affiliations. For instance, when an author publishes a document with affiliations from two countries, the document is considered for both countries. Similarly, if an article is published by the collaboration of three Iranian authors and one American author, it is counted once for both Iran and the United States.

To facilitate descriptive comparisons, we extracted the publication, citation, and collaboration bibliometric indices for article sets from HIC, LMIC, and all countries combined across pre- and post-COVID-19 periods and also for article sets from each WHO region. The complete list of the mentioned indices is provided in the suppplementary file.

Field-Weighted Citation Impact (FWCI) was considered a measure of the quality and impact of the scientific outputs. This metric is used for benchmarking purposes. FWCI is calculated by dividing the actual number of citations received by an output during the publication year plus the following three years by the total citations expected for outputs of the same type, published in the same year, and within the same journal category (43). As an instance, an FWCI of 1.45 indicates that the output is 45% more cited than expected according to its counterparts.

### 2.8 Socioeconomic indicators

To assess the relationship between socioeconomic indicators and the number of telemedicine-related publications in each country, we extracted potentially relevant indices from the World Bank website (44,45). We categorized them into three groups: indicators identified from previous studies (17,27,46,47), infrastructure-related, and telemedicine demand-related indicators. The complete list and detailed explanation of these three categories and their indices are provided in the supplementary file.

All socioeconomic indicators were extracted by year for all countries from 2018 to 2022. The average of each index was calculated using only the years with available data. These averages were used as metadata to examine the correlation between the indicators and the number of publications. We expect hospital beds, population density, the number of nurses and midwives, the number of physicians, and the percentage of the urban population residing in the largest cities to exhibit inverse relationships, while all other indicators are anticipated to demonstrate direct relationships with the general patterns in the number of publications.

### 2.9 Data extraction and preparation of data sheets

The Quacquarelli Symonds (QS) World University Rankings is a global ranking system incorporating bibliometric data from Scopus as part of its methodology for evaluating universities and institutions worldwide. It assesses university performance across five main subject areas, one of which is medicine and life sciences (48). We utilized this classification in SciVal to extract the number of telemedicine-related publications within the medicine and life sciences category.

The number of publications identified in our search for each country (Tele Pub All), the number of publications in our search in the medicine and life sciences subject area for each country (Tele Pub QS), and the total number of publications in the medicine and life sciences subject area for each country (Med Pub QS) during 2018-2022 were all exported from SciVal.

Focusing exclusively on the number of publications to assess the correlation between socioeconomic indicators and telemedicine-related scientific production can be misleading, as countries with higher overall research output are naturally more likely to produce more telemedicine-related articles. To solve this misconception and assess countries’ real interest in telemedicine research, a Research Interest (RI) index was calculated for each country (Adjusted Pub QS). This index was derived by dividing the number of telemedicine-related publications in the medicine and life sciences subject area in our search (Tele Pub QS) by the total number of publications in the medicine and life sciences subject area (Med Pub QS) during 2018–2022. Researchers often normalized the number of publications in previous studies by dividing it by a country’s population or Gross Domestic Product (GDP). This method aimed to control for the influence of a country’s overall publication volume and population size on the output in specific fields (26,27). Along with the number of publications, the correlation of RI with socioeconomic indicators was also analyzed.

Considering the volume of data and indices, we used Python software (version 3.9.13) to extract the socioeconomic indicators data, number of articles, and RI for each country between 2018-2022 and write them in datasheets against countries’ names. We collected data once for all countries and once separately for HIC and LMIC. Additionally, to compare the volume and quality of telemedicine-related scientific production between LMIC and HIC and to examine the changes in publication trends before and after the onset of COVID-19, the following additional indices were extracted from SciVal:

- Number of all telemedicine-related publications (Tele Pub All), number of telemedicine-related publications in the medicine and life sciences subject area (Tele Pub QS), and all publications in the medicine and life sciences subject area (Med Pub QS) for each country during 2018-2019 and 2020-2022
- FWCI of all telemedicine-related publications (Tele FWCI All), telemedicine-related publications in the medicine and life sciences (Tele FWCI QS), and all published papers in the medicine and life sciences subject area (Med FWCI QS) for each country during 2018-2022, 2018-2019, and 2020-2022

### 2.10 Statistical analysis

Owing to the non-normal distribution of most of the variables, we used the Spearman Rank Correlation test to assess the correlation between the mentioned socioeconomic indicators and the absolute number of articles or the adjusted number of articles (RI), categorized by HIC and LMIC. The interpretation of the correlation coefficient(r) was as follows: an r value below 0.16 was considered negligible, values between 0.16 and 0.29 indicated a weak to low relationship, values between 0.3 and 0.49 reflected a moderate to low correlation, values between 0.5 and 0.69 represented a moderate correlation, values between 0.7 to 0.89 indicated a strong relationship and values between 0.9 and 1 were classified as very strong correlations (49). To accurately assess the relationship between telemedicine-related scientific output in countries and selected socioeconomic indicators while minimizing bias, only countries with at least five publications in our dataset were included in the correlation analysis. Consequently, we analyzed data from 58 LMIC and 47 HIC.

Furthermore, given the non-normal distribution of the data, we used the Mann-Whitney U test to compare Tele Pub All, Tele Pub QS, Med Pub QS, Adjusted Pub QS, Tele FWCI All, Tele FWCI QS, and Med FWCI QS between HIC and LMIC and Wilcoxon Signed Rank test to compare the referenced indices before and after the onset of COVID-19 pandemic. Cliff’s Delta was calculated and reported with a 95% confidence interval as the effect size for these comparisons (50,51).

We applied the phasic Technique for Order Preference by Similarity to Ideal Solution (TOPSIS), a multi-criteria decision-making method to evaluate and rank WHO regions based on six selected bibliometric indices. For quality-based ranking, four quality-related indices **-** FWCI, citations per publication, the proportion of publications in the top 10% citation percentile, and the proportion of publications in the top 10% journal percentile **-** were assigned equal weights. Subsequently, the output scores of the quality-based ranking were used as inputs for a second TOPSIS model to rank the regions overall. This overall ranking incorporated two additional metrics: scholarly output as a measure of quantity and international collaboration. The weights for the overall ranking were distributed as follows: 0.4 for quality, 0.4 for scholarly output, and 0.2 for international collaboration. A two-sided p-value < 0.05 was determined as the threshold for statistical significance. All analyses were conducted using R software (version 4.3.2).

## 3. Results

### 3.1 Overview

A total of 16584 articles related to telemedicine were retrieved, including articles published between January 1, 2018, and December 30, 2022. These articles were authored by 72525 individuals and had received 282865 citations as of August 9, 2024, with an average of approximately 17 citations per publication.

The articles were imported into SciVal for analysis, and three encountered import issues. In the search limited to LMIC and HIC, 4244 documents were found for LMIC, and 13622 documents were found for HIC, including 1282 collaborative documents between the two groups. These collaborative publications accounted for 30% of LMIC outputs and 9.4% of HIC outputs.

### 3.2 Descriptive Comparison of Bibliometric Indices Between HIC and LMIC Across Pre- and Post-COVID-19 Periods

Fig 1 illustrates a comparative analysis of normalized values of bibliometric indicators between HIC and LMIC during the entire study period, as well as before and after the onset of the COVID-19 pandemic. Table S2 in the supplementary file consists of each indicator’s exact values across all income-levels and timeframes.

**Fig 1.**
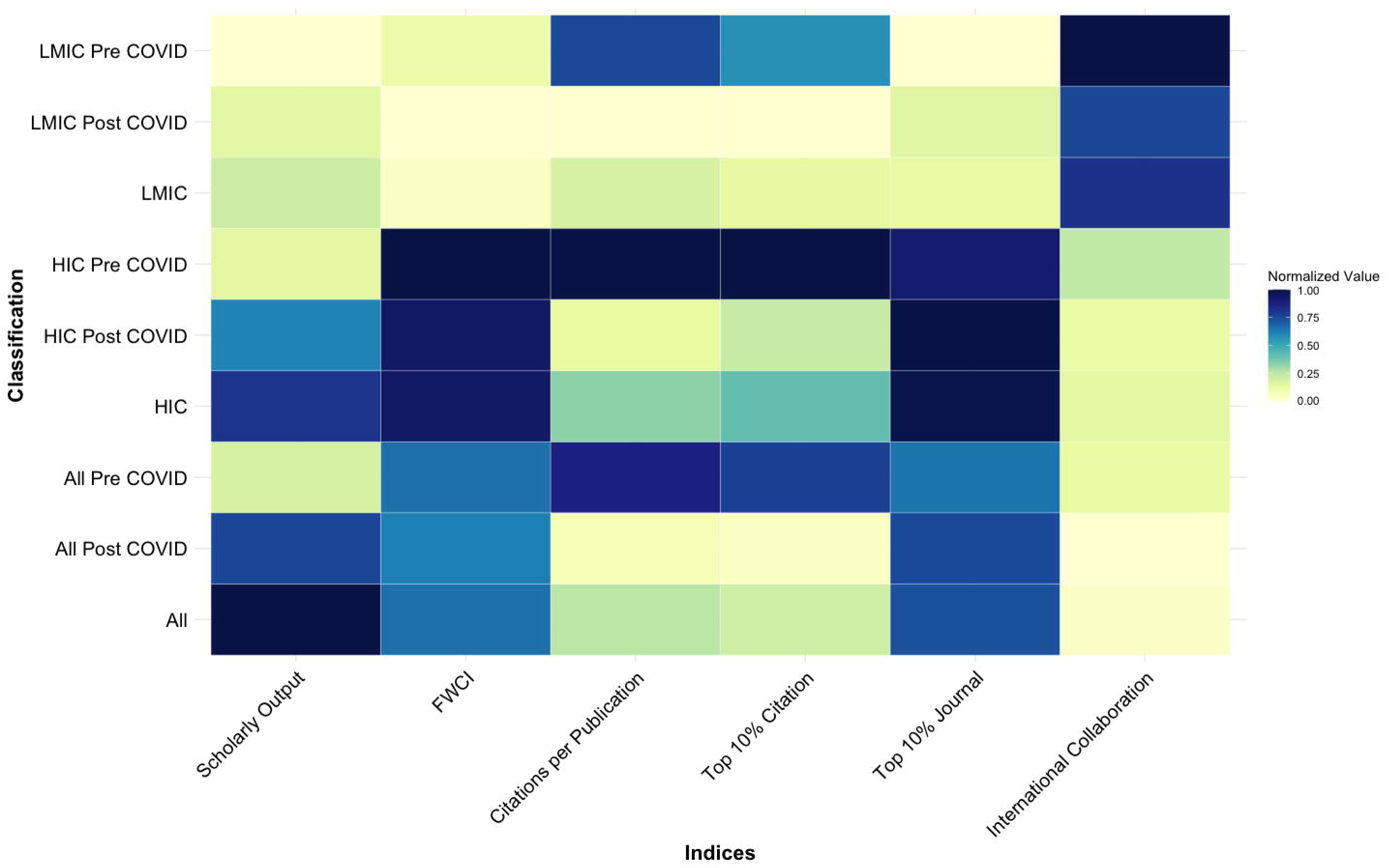
Heatmap of Normalized Telemedicine Research Bibliometric Indices Across Income and Time-Based Classifications. LMIC, Lower-Middle Income Countries; HIC, High-Income Countries; FWCI, Field-Weighted Citation Impact

#### 3.2.1 Comparison between LMIC and HIC

Scholarly outputs, citations per publication, FWCI, the proportion of outputs in the top 10% citation percentile, and the proportion of outputs in the top 10% journal percentile were consistently greater in HIC compared to LMIC across all examined timeframes. In contrast, LMIC exceeded HIC only in international collaboration, maintaining this trend in the overall period, as well as in the pre- and post-COVID-19 phases.

#### 3.2.2 Comparison between Pre- and Post-COVID-19

Scholarly outputs and the proportion of publications in the top 10% of journals increased following the onset of the COVID-19 pandemic. However, indices such as FWCI, the proportion of outputs in the top 10% of citation percentile, international collaborations, and citations per publication decreased post-COVID-19 across all groups, including LMIC and HIC. It is important to note that citations per publication and the proportion of articles in the top 10% citation percentile are inherently influenced by the time elapsed since publication. Consequently, direct comparisons of these metrics between pre- and post-COVID-19 periods may not be entirely reliable

### 3.3 Analytical comparison of the volume and FWCI of telemedicine-related publications between HIC and LMIC Across Pre- and Post-COVID-19 Periods

Fig 2 provides the effect sizes with 95% Confidence Intervals for tests comparing publication volume-related metrics (Tele Pub All, Tele Pub QS, Med Pub QS), Adjusted Pub QS (RI), and publication quality-related metrics (Tele FWCI All, Tele FWCI QS, and Med FWCI QS) across different timeframes and income groups. We did not compare publication volumes between the pre- and post-COVID periods, as the pre-COVID period included articles from two years, while the post-COVID period covered three years, making direct comparison unreliable and inaccurate. Moreover, Effect size categories were determined based solely on point estimates. However, the majority of confidence intervals spanned two or more effect size categories, rendering many of the comparisons relatively inconclusive. Table S3 in the supplementary file provides detailed results of these comparisons.

**Fig 2.**
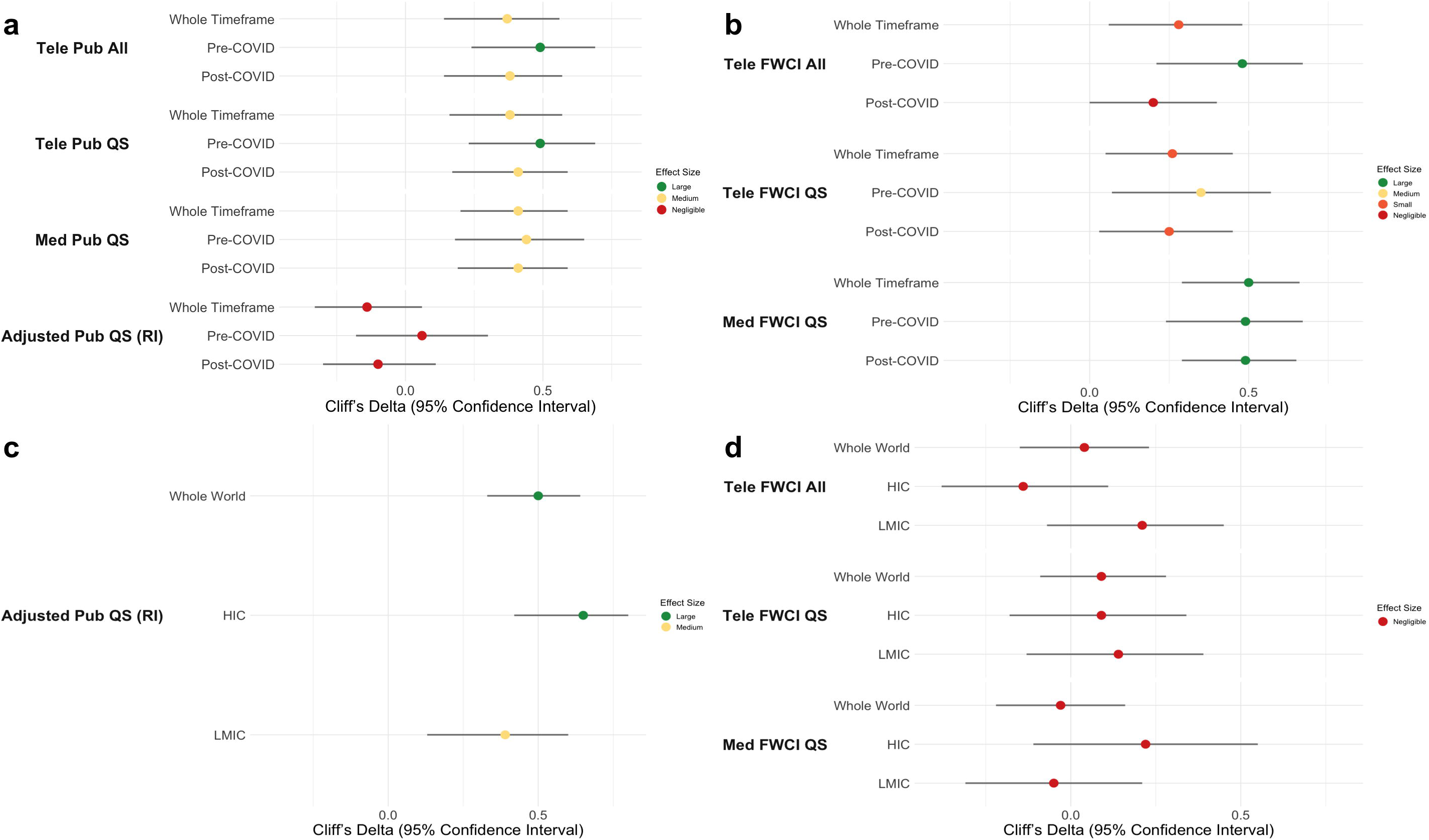
Effect Size (Cliff’s Delta) for Comparisons of Selected Bibliometric Indices. (a) Comparisons of Volume-related Indices and RI Between HIC and LMIC in Given Timeframes. (b) Comparisons of Quality-related Indices (FWCI) Between HIC and LMIC in Given Timeframes. (c) Comparisons of RI Between Pre- and Post-COVID Periods in Given Group of Countries. (d) Comparisons of Quality-related Indices (FWCI) Between Pre- and Post-COVID Periods in Given Group of Countries. LMIC, Lower-Middle Income Countries; HIC, High-Income Countries; FWCI, Field-Weighted Citation Impact; RI, Research Interest; QS, Quacquarelli Symonds

#### 3.3.1 Comparison between LMIC and HIC

As illustrated in Fig 2, the number of telemedicine-related publications across all subject areas (Tele Pub All) and those within the medicine and life sciences subject area (Tele Pub QS) were significantly higher in HIC compared to LMIC across all timeframes. These differences were more prominent in the pre-COVID era (both Cliff’s Delta = 0.49)(Fig 2a)

HIC also produced a significantly higher number of publications in the medicine and life sciences subject area (Med Pub QS) overall compared to LMIC across all timeframes. Medium effect sizes were observed across all timeframes, with the largest difference noted in the pre-COVID era (Cliff’s Delta = 0.44). Interestingly, RI did not differ significantly between HIC and LMIC within any timeframe.(Fig 2a)

The FWCI of total articles in the medicine and life sciences subject area (Med FWCI QS) was significantly higher in HIC compared to LMIC in all timeframes. The effect sizes for all comparisons fell within the large range, indicating substantial differences. (Fig 2b)

The quality and impact of the telemedicine-related publications in the medicine and life sciences subject area (Tele FWCI QS) was significantly higher in HIC than in LMIC, with the largest difference observed in the pre-COVID era (Cliff’s Delta = 0.35). (Fig 2b)

However, when comparing the FWCI of telemedicine-related publications overall (Tele FWCI All), no statistically significant difference was seen in the post-COVID era. A small difference was noted during the entire study timeframe (Cliff’s Delta = 0.28), while a large difference was evident in the pre-COVID era (Cliff’s Delta = 0.48). (Fig 2b)

#### 3.3.2 Comparison between Pre- and Post-COVID-19 periods

The onset of the COVID-19 pandemic resulted in a significant increase in RI (Adjusted Pub QS), with large effect sizes observed across all countries and HIC and a medium effect size in LMIC. Additionally, the RI in telemedicine increased more significantly in HIC compared to LMIC (Cliff’s Delta = 0.65 vs Cliff’s Delta = 0.39). (Fig 2c)

There were no statistically significant differences between publication quality-related metrics after the onset of the COVID-19 pandemic in all countries, HIC and LMIC. (Fig 2d)

### 3.4 Descriptive comparison of bibliometric indices between WHO Regions

The U.S. publications heavily influenced the number of publications in the Americas region. Therefore, we analyzed and presented the data both, including and excluding U.S. publications (Fig 3), to provide a clearer perspective. The chart highlights regional disparities in research quantity and quality. Notably, these comparisons are solely based on the magnitude of the indices and do not imply statistical significance. Table S4 in the supplementary file shows each indicator’s exact values across all WHO regions.

**Fig 3.**
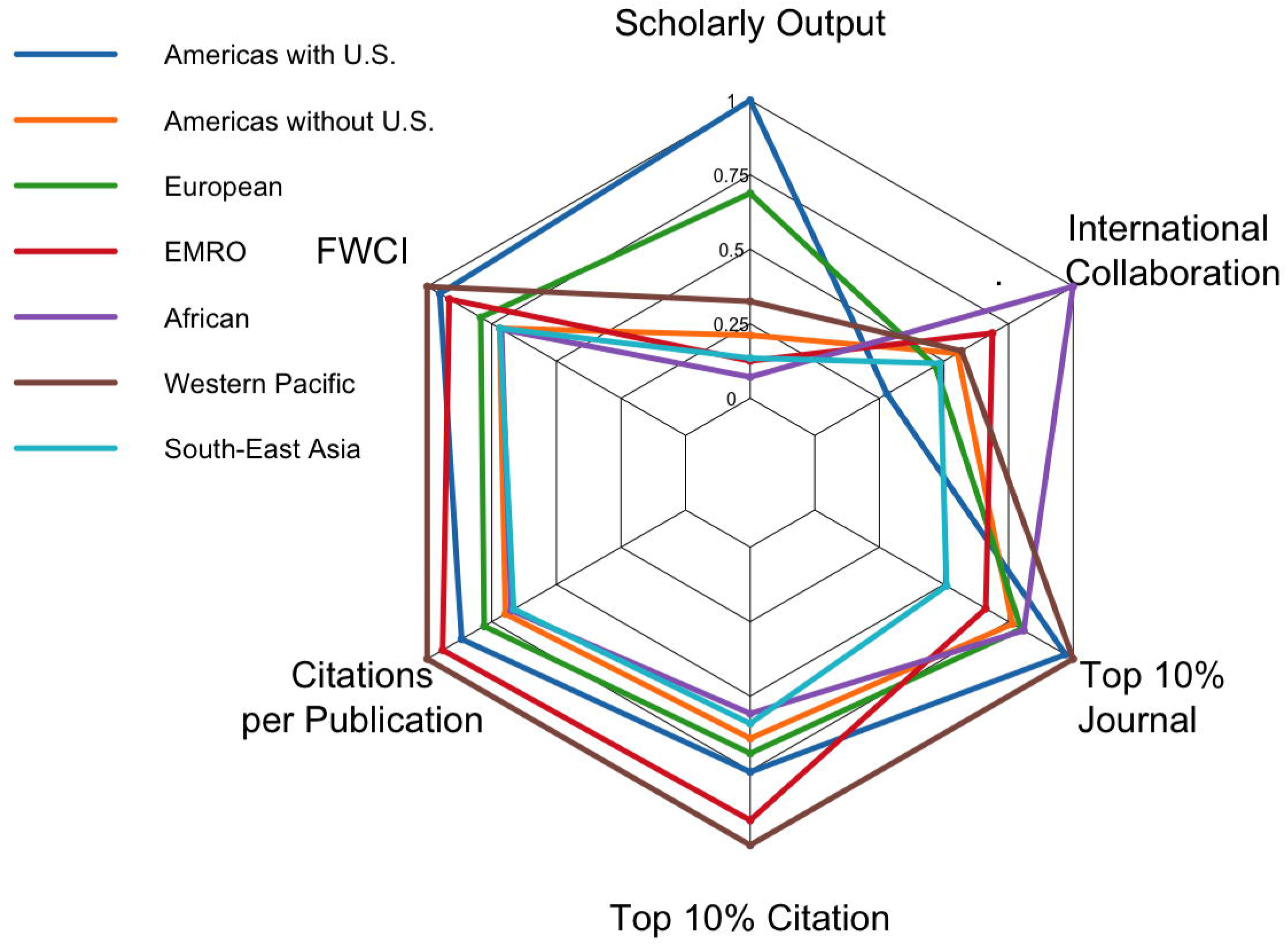
Spider chart illustrating the comparative performance of World Health Organization (WHO) regions across selected bibliometric indices, including Scholarly Output, Field-Weighted Citation Impact (FWCI), Citations per Publication, Top 10% Citation Percentile, Top 10% Journal Percentile, and International Collaboration. U.S., United States of America; EMRO, Eastern Mediterranean Region

When comparing WHO regions, the Americas region published the highest number of telemedicine-related articles, owing to contributions from the U.S. and Canada, followed by the European region. Conversely, the African region had the fewest publications, with the Eastern Mediterranean region (EMRO) ranking second lowest.

The highest FWCI among WHO regions was observed in the Western Pacific, indicating the high quality of telemedicine-related publications in this region. In contrast, the African region had the lowest FWCI. Notably, EMRO’s FWCI surpassed that of the European region. However, when the U.S. was excluded, the quality of telemedicine-related publications from the Americas region declined to levels in alignment with those of the African and Southeast Asian regions.

The Western Pacific region ranked first when comparing the citations per publication index across WHO regions. The EMRO followed as the second rank, aligning with its third-place ranking in FWCI. The Americas region ranked third in the citations per publications index. As expected, the lowest values of this index were observed in the Southeast Asian and African countries, respectively.

The international collaboration rate was highest among publications of the African region and lowest among those from the Americas region. Following African countries, we observed the second-highest level of international collaboration in EMRO, with approximately half of its publications involving international partnerships. On the other hand, the Americas, European, and Southeast Asian regions showed the lowest rates of international collaboration.

The proportion of publications in the top 10% citation percentile is ranked similarly to the FWCI ranking, with the Western Pacific region leading and the African region at the bottom.

Finally, researchers from the Western Pacific region published the highest proportion of publications in the top 10% journal percentile, followed by the Americas region. Despite a strong performance by the EMRO in other quality metrics, such as FWCI, citations per publication, and outputs in the top 10% citation percentile, researchers from this region published the fewest telemedicine-related articles in top-tier journals, second only to Southeast Asia.

Using the TOPSIS method, we proposed quality-based and overall rankings of WHO regions according to previously mentioned indices of their telemedicine-related publications (Fig 4). The Southeast Asia region ranked lowest in both quality and overall performance. The Western Pacific region ranked highest in quality but came second overall. The Americas region, when including the U.S., ranked highest overall but second to the Western Pacific region in quality. The EMRO region outperformed the European region in quality but not in overall ranking, where Europe placed third. Excluding the U.S., The Americas region ranked even lower than the African region in both quality and overall rankings, placing last after the Southeast Asia region. Detailed scores from the TOPSIS analyses for both ranking processes are provided in the supplementary file (Table S5 and S6).

**Fig 4.**
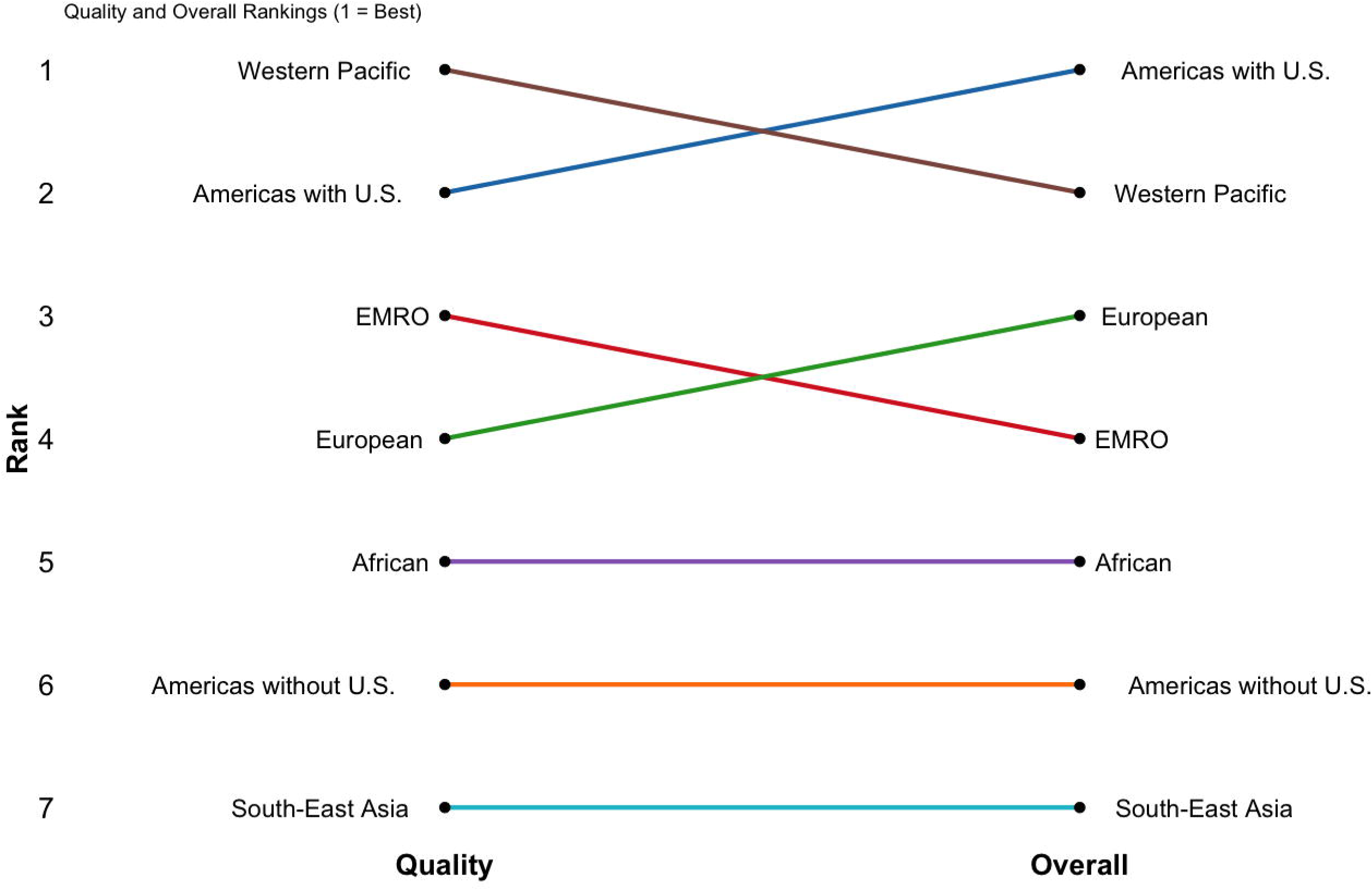
Comparison of WHO region rankings based on quality-specific and overall bibliometric performance. Rankings are displayed on a reversed scale (1 = best). U.S., United States of America; EMRO, Eastern Mediterranean Region

Lastly, Iran underperformed in quality-related indices of telemedicine research compared to the averages for EMRO countries and the global standard despite publishing the highest number of articles among EMRO countries. (Table S4).

### 3.5 Socioeconomic Drivers

We examined the correlations between the selected socioeconomic indicators and telemedicine-related output from two perspectives: the absolute number of publications and RI. Table S7 in the supplementary file presents the correlation coefficients and p-values from correlation tests performed separately for HIC and LMIC. Additionally, the Table shows the strength and direction of statistically significant relationships between the indicators and publication output measures (the absolute number of articles and RI). Table S8 in the supplementary file classifies the indicators based on their observed relationships, separated by HIC and LMIC. Fig 5 depicts the most influential factors affecting telemedicine research production in both HIC and LMIC. A detailed explanation and evaluation of these correlations are provided in the discussion section.

**Fig 5.**
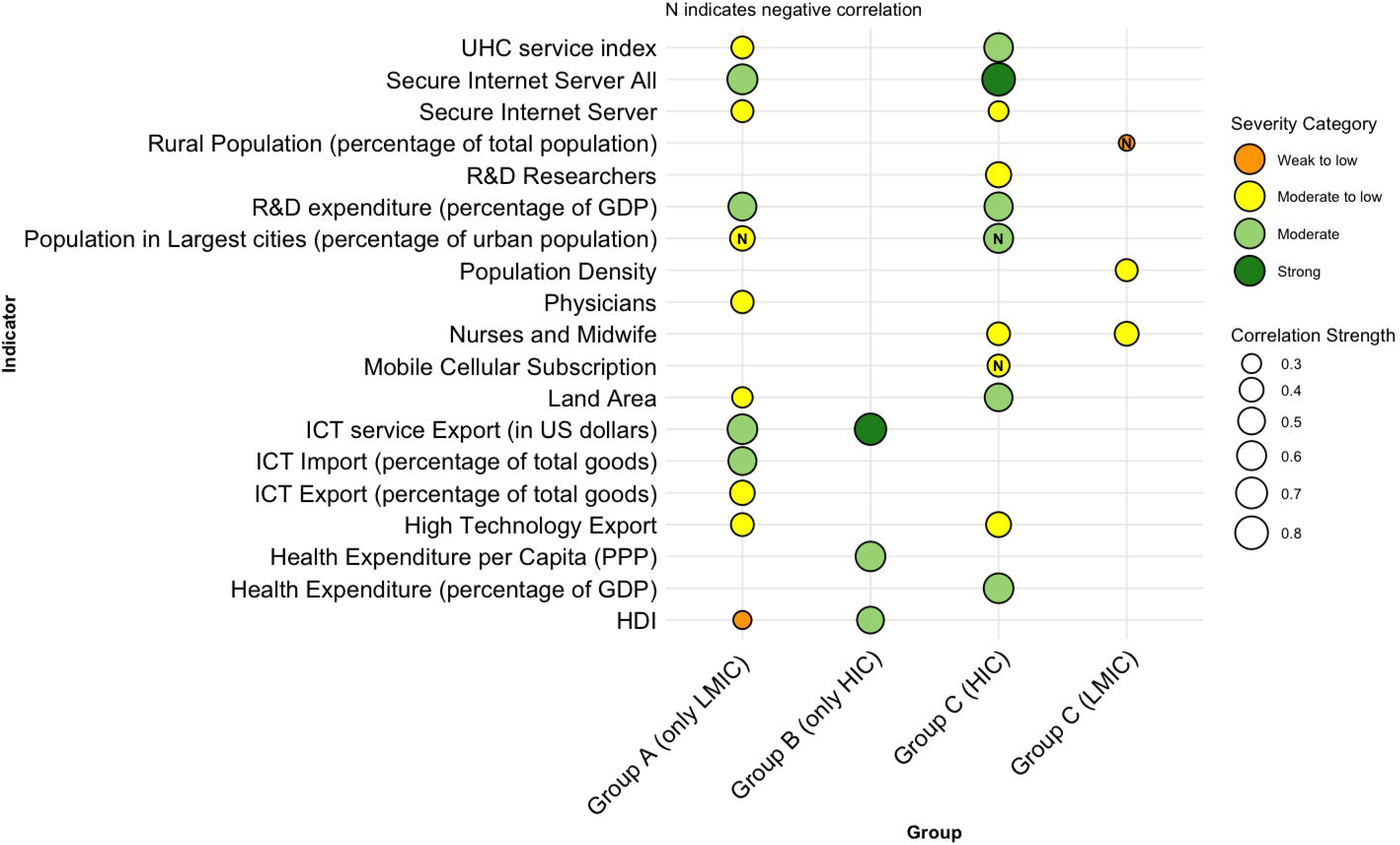
Bubble chart illustrating the socioeconomic drivers potentially associated with telemedicine scientific output, as identified in the present study. The size of each bubble reflects the strength of the correlation, while the presence of the letter ’N’ denotes a negative association. Severity categories are color-coded and reflect the magnitude of correlation. Group D is excluded from the figure due to the likelihood of incidental findings, and Group E is omitted as no meaningful correlation was observed—details discussed in the main text. Associations are based on correlation with the absolute number of telemedicine-related publications. UHC, Universal Health Coverage; R&D, Research and Development; GDP, Gross Domestic Product; ICT, Information and Communication Technologies; US, United States; PPP, Purchasing Power Parity; HDI, Human Development Index

**Group A** comprises indicators correlated with both the absolute number of telemedicine-related articles and RI in telemedicine but in the opposite directions (these patterns were only detected in LMIC). Among the indicators, all correlations were in the expected direction except for the number of physicians. It was expected that a higher rate of physicians per population would lead to a reduced focus on telemedicine; nevertheless, the data revealed the opposite.

**Group B** includes indicators correlated with both the absolute number of telemedicine-related articles and RI in telemedicine in the same direction (these patterns were only detected in HIC).

**Group C** consists of indicators correlated only with the absolute number of telemedicine-related articles. Notably, in LMIC, the direction of all correlations was contrary to expectations. Conversely, in HIC, all indicators were correlated in the anticipated direction, except the number of nurses and midwives and mobile cellular subscriptions. The former was expected to have an inverse relationship with research output, while the latter was anticipated to correlate positively with telemedicine-related research output.

**Group D** encompasses indicators that were correlated only with RI in telemedicine. In LMIC, all the correlations were contrary to expectations, while in HIC, the correlations of Hospital Beds and percentage of population Using Internet with RI were consistent with expectations, whereas those of percentage of population aged 65 and above and percentage of Rural population were in the opposite direction.

**Group E** comprises indicators which were not correlated with neither the absolute number of telemedicine-related articles nor telemedicine RI.

## 4. Discussion

HIC outperformed LMIC across multiple bibliometric indices, publishing approximately three times more telemedicine-related articles. This disparity was consistent across all timeframes. However, the research output gap between LMIC and HIC has narrowed since the COVID-19 pandemic, suggesting that the pandemic may have contributed to the convergence of research productivity between these regions. Notably, the lag in research output among LMIC is not unique to telemedicine but is observed across most scientific fields. (17–19,52,53). Some research outputs from LMIC are not indexed in major international databases, further underestimating their contribution to the field (54). Researchers in LMIC face significant challenges, including limited research funding, language barriers, and the high cost of publishing in prestigious journals, all of which may contribute to their lower research output. (55). The trends in the number of telemedicine articles appear to reflect broader patterns in global research activity, suggesting that these trends are not exclusive to telemedicine but are derived from systemic inequalities in scientific output (25,56).

Interestingly, the RI in telemedicine did not differ significantly between HIC and LMIC across all timeframes. This finding suggests that the greater number of telemedicine-related publications from HIC is primarily due to their overall larger research output rather than a specific emphasis on telemedicine. Additionally, the RI for telemedicine increased significantly following the emergence of the COVID-19 pandemic, indicating that the surge in telemedicine-related publications was not merely a byproduct of the overall rise in research output but rather a reflection of telemedicine becoming a higher research priority. Notably, this increase in RI was more pronounced in HIC than in LMIC.

Comparing HIC and LMIC regarding impact metric (FWCI), a significant difference was observed in telemedicine-related publications within the medical subgroup across all timeframes. Although the effect size and intensity were smaller than those seen for the broader medical category, HIC consistently outperformed LMIC. This finding suggests that telemedicine research may offer a more equitable platform for LMIC to produce impactful contributions, helping to narrow the gap in scholarly influence compared to other areas of medical research. When analyzing the FWCI of all telemedicine-related publications, no statistically significant difference was observed between HIC and LMIC in the post-COVID-19 period. However, a substantial difference was noted in the pre-COVID-19 era. These findings suggest that the COVID-19 pandemic may have contributed to a convergence in the impact and quality of telemedicine-related research between HIC and LMIC.

LMIC exhibited a higher rate of international collaboration than HIC, a pattern that remained consistent across all timeframes. In LMIC, approximately 4 out of every 10 publications involved international collaboration, with nearly three of these four including at least one contributor from a high-income country. In contrast, only 2 out of every 10 articles affiliated with HIC had an international collaborator, with half of these involving at least one contributor from a low-income country. This may highlight the substantially higher proportion of HIC contributions to LMIC-affiliated publications compared to the reverse (three vs. one out of every 10 articles, respectively). It also suggests the potentially greater influence of HIC authors on LMIC research in the field of telemedicine. This pattern may be attributed to research in LMIC being predominantly shaped by the priorities and interests of HIC, potentially overlooking the specific healthcare needs of LMIC populations (57).

There are discrepancies within the research output and quality metrics among different WHO regions. While EMRO ranked fifth out of six in telemedicine scientific output, its FWCI and citations per publication (impact and quality-related metrics) ranked third and second, respectively. This indicates that EMRO produced high-quality telemedicine articles despite its low publication volume. Moreover, the findings suggest that the high quality and impact of EMRO publications were driven by countries other than Iran. We observed a similar trend in the Western Pacific region, which, although producing fewer articles compared to the Americas and European regions, ranked first in impact-related metrics. This is probably due to studies conducted in Japan, China, South Korea, and particularly Australia. In contrast, the African and Southeast Asian regions performed the worst among WHO regions regarding scholarly outputs, FWCI, and citations. Additionally, our findings highlight the dominant role of the United States in telemedicine research production within the Americas region. Excluding the U.S., the impact and volume of telemedicine research in this region ranked second to last, only ahead of Southeast Asia.

Group A correlations were observed exclusively among LMIC. As detailed in the supplementary file (Fig S2), the ranking of countries by the number of telemedicine publications among LMIC was relatively inverse to their ranking by RI. This suggests that countries producing a higher volume of telemedicine publications may have produced even more overall medical publications, resulting in this reverse direction. These countries seem to have allocated a smaller proportion of their research efforts to telemedicine (as reflected in RI). However, this inverse relationship was not evident among HIC, where RI and absolute number of publication rankings did not follow a particular pattern (Fig S1 in the supplementary file). Among these socioeconomic drivers, the number of physicians was the only indicator correlated with absolute numbers contrary to expectations. This finding suggests that countries with more physicians may possess better infrastructure for conducting research and implementing telemedicine, even if telemedicine prominence is expected to rise in contexts with fewer physicians. Similarly, one study observed a positive correlation with the number of healthcare practitioners per 1000 inhabitants by analyzing telemedicine-related publications from 1964 to 2003 (26). Research and Development (R&D) expenditure as a percentage of GDP (r = 0.55), the total number of secure internet servers (r = 0.65), ICT imports as a percentage of total imports (r = 0.55), and ICT service exports in dollar value (r = 0.63) were the most prominent associated factors with telemedicine scientific production in LMIC (Fig 5).

Group B indicators and their associations were observed exclusively among HIC. Health Expenditure per capita adjusted for Purchasing Power Parity (PPP) (r = 0.63, r = 0.45), HDI (r = 0.5, r = 0.47), and ICT service exports in dollar value (r = 0.72, r = 0.33) were three indicators with the highest probability of being positively correlated with both telemedicine scientific output and RI in HIC (Fig 5). Likewise, another study reported a positive correlation between the HDI and the number of teleophthalmology publications per million population (27). Similarly, another study reported a positive correlation between teledermatology and telemedicine scientific output across 60 countries and socioeconomic indicators such as GDP, GDP per hour worked, and HDI(17). In contrast, one study identified a negative correlation between telemedicine publications per capita and HDI. Moreover, they reported a positive correlation with GNP per capita (26). Differences in the studies’ timeframes and methods of scientific output normalization may result in variations in findings.

The lack of correlation with RI despite correlation with absolute numbers among group C indicators may suggest either two possibilities: first, the observed relationships may be attributed to the overall research output of these countries rather than being specific to telemedicine; Second, the observed relationships could be real, and these indicators only correlated with the absolute number of telemedicine publications, not with telemedicine-specific research interest. In HIC, the total Secure Internet Servers was strongly positively correlated (r = 0.8) with the absolute number of publications, While Health Expenditure as a Percentage of GDP (r = 0.66), R&D Expenditure as a Percentage of GDP (r = 0.56), Land Area (r = 0.54), Universal Health Coverage (UHC) service index (r = 0.58), and Percentage of Urban Population living in Largest Cities (r = 0.6) had a moderate positive correlation. The remaining indicators demonstrated moderate-to-weak correlations. These factors, along with group B indicators, might be the most likely socioeconomic drivers of telemedicine scientific production in HIC, confirmed by previous studies (17,26).

Group A and C indicators may correlate with scientific production overall, not specifically with telemedicine production, while Group B indicators warrant a stronger relationship with telemedicine production.

Group D relationships could be incidental and reflective of the higher heterogeneity in the total number of medical publications compared to the relatively homogeneous distribution of telemedicine-related publications. This heterogeneity might result in correlations with RI but not with the absolute number of publications, limiting the interpretive value of findings, even in HIC. Most of these associations were observed in LMIC with low intensity and in the opposite direction of what was expected, suggesting their accidental nature and limited importance. Lastly, Group E indicators probably have no significant relationship with scientific production in telemedicine or even with overall scientific production.

The relationship between socioeconomic indicators and bibliometric indices has been extensively studied in fields beyond telemedicine as well, presenting mixed and contradictory results summarized in Table S9 (21–25,46,58–73). Studies investigating socioeconomic drivers in the telemedicine research field are also presented in Table 1. These findings suggest that several of the observed associations are not unique to telemedicine but reflect broader trends in a country’s overall scientific production. Moreover, the strength of the relationship between socioeconomic factors and the volume of publications on telemedicine was weaker in this study compared to findings from other fields and other telemedicine studies (71,72).

**Table 1.**
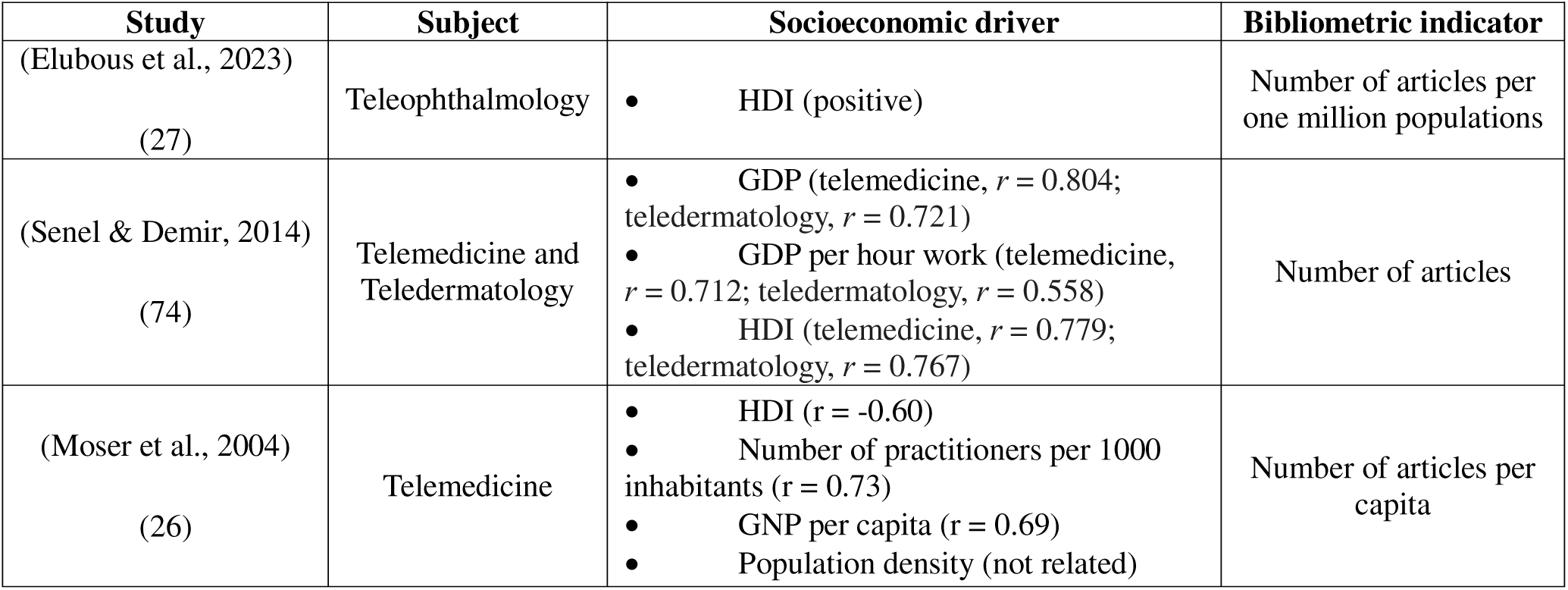
Similar studies investigating socioeconomic drivers in telemedicine research field.

| Study | Subject | Socioeconomic driver | Bibliometric indicator |
| --- | --- | --- | --- |
| (Elubous et al., 2023)<br>(27) | Teleophthalmology | HDI (positive) | Number of articles per one million populations |
| (Senel & Demir, 2014)<br>(74) | Telemedicine and Teledermatology | GDP (telemedicine, $r = 0.804$; teledermatology, $r = 0.721$)<br>GDP per hour work (telemedicine, $r = 0.712$; teledermatology, $r = 0.558$)<br>HDI (telemedicine, $r = 0.779$; teledermatology, $r = 0.767$) | Number of articles |
| (Moser et al., 2004)<br>(26) | Telemedicine | HDI ($r = -0.60$)<br>Number of practitioners per 1000 inhabitants ($r = 0.73$)<br>GNP per capita ($r = 0.69$)<br>Population density (not related) | Number of articles per capita |

The pattern of correlations between socioeconomic factors and research output in HIC compared to LMIC was relatively consistent with the existing literature. Other studies demonstrated that the socioeconomic factors in HIC have predominantly correlated with scientific production (67). In contrast, these factors in LMIC were assessed in a few mentioned studies (63,69), either showing no correlation or demonstrating unexpected relationships with scientific output without any particular trends.

## Limitations and Suggestions for future work

The present study is not without limitations. First, the results were derived exclusively from a single database, Scopus. While Scopus is among the most comprehensive databases, particularly for publications from LMIC, utilizing multiple databases could provide a more comprehensive understanding of trends and mitigate potential biases. Second, our timeframe for analysis was relatively short. Extending the study period in future research would allow for a more comprehensive evaluation of telemedicine-related publications, providing greater insights into trends and changes over time. This would also enhance the generalizability of the findings.

Third, the evaluation of the relationship between socioeconomic indicators and telemedicine-related research output is multifactorial, involving numerous intermediary and confounding variables. Additionally, collinearity among many of these indicators complicates interpretation. Therefore, conclusions regarding these relationships need to be drawn with caution. Our study focused solely on assessing correlations between these variables from a univariate perspective. Future research is encouraged to employ multivariate analyses to account for the interactions among indicators and more accurately estimate each indicator’s independent effects on research output.

Fourth, we used the screening method described in the Methods section as an alternative to the classical approach of title and abstract review followed by full-text review. While more efficient, this method is inherently less precise than the classical approach and may introduce bias into the final results.

Fifth, the exclusion of telemedicine-related publications in subject areas beyond medicine and life sciences from the RI indicators would slightly skew the results. Since RI is solely a measure of research interest within medicine and life sciences, this limitation should be considered when drawing conclusions. Moreover, using the FWCI as a measure of research quality may also introduce bias, as it relies only on citation counts. Since citations are only one aspect of research quality, their use as the sole metric should be interpreted cautiously.

It is good to consider two additional points in our study. First, LMIC represent a more heterogeneous group compared to HIC, encompassing a range from the world’s poorest nations to upper-middle-income and developed countries, such as China. This diversity may influence some of the study’s findings. Second, as noted, Scopus and SciVal count an article for each author’s country based on their institutional affiliation without paying attention to the extent of their contribution to the study. This limitation could hinder achieving conclusive results.

## 5. Conclusion

Global inequalities persist in telemedicine research, often favoring high-income and developed regions. A significant portion of research in LMIC is conducted in collaboration with HIC, raising concerns about the potential neglect of region-specific research needs. However, the COVID-19 pandemic may have mitigated this gap by increasing the volume and quality of telemedicine-related publications and advocating global interest in telemedicine research.

A meaningful alignment between various socioeconomic variables, research output, and research interest was observed only in HIC, depicting a more proportionate telemedicine research pattern corresponding to local research capabilities, infrastructure, and domestic needs. In this context, and exclusively within HIC, Health Expenditure as a percentage of GDP, R&D Expenditure as a percentage of GDP, Land Area, and UHC service index exhibited a moderate positive correlation with crude telemedicine research output, while Secure Internet Servers All showed a strong positive correlation. Additionally, Health Expenditure per capita (PPP) and HDI demonstrated a moderate positive correlation, whereas ICT Service Exports in the U.S. dollar exhibited a strong positive correlation with both telemedicine research output and RI.

## Data Availability

The spreadsheets containing bibliometric data and socioeconomic indicators retrieved for each country and used in the analyses are available and can be accessed upon reasonable request from the corresponding author.
Moreover, the data used in this study were obtained from Scopus and SciVal, which are subscription-based services provided by Elsevier. Access to these databases is subject to institutional licensing agreements, and the data cannot be publicly shared. Researchers interested in replicating the analyses should obtain access to Scopus and SciVal through their institutions.

## Acknowledgements

Not applicable

## Statements and Declarations

### Consent to participate

Not applicable. This study did not involve human participants, personal data or animal; it was based solely on the analysis of published articles.

## Author Contributions

S.S.A & F.S: Investigation, Methodology, Data curation, Writing-original draft/ S.S.A: Formal analysis, Visualization/ M.S: Conceptualization, supervision, Writing-review and editing. All authors have read and approved the final version of the manuscript for submission. M.S. is guarantor.

## Data availability statement

The spreadsheets containing bibliometric data and socioeconomic indicators retrieved for each country and used in the analyses are available and can be accessed upon reasonable request from the corresponding author.

Moreover, the data used in this study were obtained from Scopus and SciVal, which are subscription-based services provided by Elsevier. Access to these databases is subject to institutional licensing agreements, and the data cannot be publicly shared. Researchers interested in replicating the analyses should obtain access to Scopus and SciVal through their institutions.

## Abbreviations

LMIC: Low- and Middle-Income Countries
HIC: High-Income Countries
WHO: World Health Organization
ICT: Information and Communication Technologies
CDC: Centers for Disease Control and Prevention
HDI: Human Development Index
GNP: Gross National Product
PCP: Primary Care Providers
GNI: Gross National Income
FWCI: Field-Weighted Citation Impact
GDP: Gross Domestic Product
RI: Research Interest
TOPSIS: Technique for Order Preference by Similarity to Ideal Solution
U.S.: United States of America
EMRO: Eastern Mediterranean region
UHC: Universal Health Coverage
R&D: Research and Development

