## Supplemental Material for "The Global Imbalance in Telemedicine Research: An Analysis of Knowledge Production and Socioeconomic Drivers"

Supplementary Material

### 1 Search string and its rationale

The rationale for the search strategy:

1. Keywords related to Telemedicine and Telehealth (Title)
2. Keywords related to Telecare (Title)
3. Keywords related to Mobile health (Title)
4. Keywords related to Electronic health (Title)
5. Keywords related to Teleconsultation and e-consultation (Title)
6. Keywords related to Telerehabilitation (Title)
7. Keywords related to Telehealthcare (Title)
8. Keywords related to various medical fields (e.g., Teleradiology) (Title)
9. Keywords related to Virtual visits and Virtual clinics (Title)
10. Publication Year: 2018-2022
11. Excluded publication types: Conference papers, notes, editorials, erratum, letters, and book chapters

Final search query:

(1 OR 2 OR 3 OR 4 OR 5 OR 6 OR 7 OR 8 OR 9) AND 10 AND 11

( TITLE ( telemed* OR "tele med*" OR telehealth OR "tele health" OR telecare OR "tele care" OR "remote care" OR "video care" OR "tele intensive care" OR "tele icu" OR "mobile health" OR mhealth OR "m health" OR "health, mobile" OR ehealth OR "e health" OR ecare OR "e care" OR econsult* OR "e consult*" OR ediagnos* OR "e diagnos*" OR "electronic diagnos*" OR enurs* OR "e nurs*" OR "electronic nurs*" OR ephysician* OR "e physician*" OR epsych* OR "e psych*" OR etherap* OR "e therap*" OR "electronic therap*" OR emedicine* OR "e medicine*" OR "electronic medicine" OR eicu OR "e icu" OR teleradiolog* OR "tele radiolog*" OR telepsychiatr* OR "tele psychiatr*" OR telepsycholog* OR "tele psycholog*" OR telepatholog* OR "tele patholog*" OR teledermatolog* OR "tele dermatolog*" OR telegenetic* OR "tele genetic*" OR teleophthalmolog* OR "tele ophthalmolog*" OR telepharmac* OR "tele pharmac*" OR teleemergenc* OR "tele emergenc*" OR telesurge* OR "tele surge*" OR teleradiotherap* OR "tele radiotherap*" OR teleretin* OR telestroke OR "tele stroke" OR telenurs* OR "tele nurs*" OR "telemental health" OR teledent* OR "tele dent*" OR telerahab* OR "tele rehab*" OR "telediabete*" OR "tele diabete*" OR "remote rehab*" OR "virtual rehab*" OR "home rehab*" OR "rehab*, remote" OR "rehab*, virtual" OR econsult* OR "e consult*" OR "electronic consult*" OR "online consult*" OR teleconsult* OR "tele consult*" OR "remote consult*" OR "web consult*" OR videoconsult* OR "video consult*" OR "virtual consult*" OR "telephone consult*" OR ( remote W/2 ( consult* OR visit* ) ) OR ( email W/2 ( consult* OR appoint* ) ) OR ( text W/2 ( consult* OR appoint* ) ) OR "consult*, remote" OR "web counsel*" OR "remote counsel*" OR "distance counsel*" OR "video counsel*" OR telecounsel* OR "tele counsel*" OR teleclinic* OR "tele clinic*" OR "remote clinic*" OR "video clinic*" OR telehomecare OR "tele home care" OR telehealthcare OR "tele healthcare" OR "remote health*" OR "virtual health*" OR "video visit*" OR "virtual visit*" OR "digital psychological intervention" OR "mental health teletherapy" OR "remote medicine" OR "virtual medicine" OR "medicine, virtual" OR teletreat* OR "tele treat*" OR telefollow* OR "tele follow*" OR telecure OR "tele cure" OR teletherap* OR "tele therap*" OR telediagno* OR "tele diagno*" OR "tele refer*" OR telerefer* OR teleprocedu* OR "tele procedu*" OR teleinterven* OR "tele interven*" OR telepractic* OR "tele practic*" OR telebehav* OR "tele bahav*" ) OR ( TITLE ( telecommunicat* OR "tele communicat*" OR telematic* OR "tele matic*" OR teleconferenc* OR "tele conferenc*" OR teleassist* OR "tele assist*" OR teleservice* OR "tele service*" OR telemanag* OR "tele manag*" OR telescreen* OR "tele screen*" OR telemonitor* OR "tele monitor" OR "telephone monitor*" ) AND ( SUBJAREA ( medi OR nurs OR vete OR dent OR heal OR mult ) ) ) ) AND PUBYEAR > 2017 AND PUBYEAR < 2023

### 2 Bibliometric Indices

- Publication metrics
  - Number of documents (scholarly output)
  - Proportion of outputs in the top 10% citation percentile
  - Proportion of outputs in the top 10% journal percentile
- Citation metrics
  - Field Weighted Citation Impact (FWCI)
  - Citations per publication
- Collaboration metrics
  - International collaboration rate

### 3 Socioeconomic Drivers

*Indicators identified from previous studies***:** some indices were chosen based on their examination in previous studies that investigated their relationship with the number of publications:

- Health Expenditure (percentage of Gross Domestic Product (GDP))
- Health Expenditure per capita (purchasing power parity (PPP)): health expenditure in US dollars adjusted for PPP
- Human Development Index (HDI)
- Research and Development (R&D) expenditure (percentage of GDP)
- R&D Researchers: Number of researchers engaged in R&D per one million people
- Population Density: Mid-year population divided by land area
- Nurses and Midwives: Number of nurses and midwives per 1000 people
- Physicians: Number of physicians per 1000 people

*Infrastructure-related indicators:* Several other indices reveal the state of a country’s infrastructure for implementing telemedicine, considered as indirect measures of telemedicine utilization capacity:

- Electricity accessibility (percentage of Rural population)
- Population Using Internet (percentage of total population)
- Secure Internet Servers: Number of secure servers per one million people, measured when a computer connects to a website server
- Secure Internet Server All: Total number of secure internet servers
- High-Technology Exports: products with high R&D intensity, such as aerospace, computers, pharmaceuticals, and electrical machinery measured in US dollars
- Mobile Cellular Subscription: Mobile cellular subscriptions per 100 people
- Literacy Rate: percentage of people aged 15 and above who can both read and write
- Information and communication technology (ICT) goods Exports (percentage of total goods): proportion of total goods exports in a country allocated to ICT goods, including computers and peripheral equipment, communication equipment, consumer electronic equipment, electronic components, and other information and technology goods
- ICT goods Imports (percentage of total goods): proportion of total goods imports in a country allocated to ICT products
- ICT service Exports (percentage of service exports): proportion of service exports involving computer and communications services (telecommunications and postal and courier services) and information services (computer data and news-related service transactions)
- ICT service Exports (in U.S. dollars): Value of ICT service exports in US dollars

*Telemedicine Demand-related indicators:* A few more indicators reflect the need for telemedicine implementation in a country, under the assumption that each country conducts telemedicine research proportionate to its demand:

- Hospital Beds: Number of inpatient beds available per 1000 people
- Land Area: Total land area of the country, excluding water bodies
- Population aged 65 and above (percentage of total population)
- Rural Population (percentage of total population)
- Population in Largest cities (percentage of urban population): percentage of a country’s urban population living in the largest metropolitan area
- Universal Health Coverage (UHC) service index: a number between 0 and 100 representing the coverage index for essential health services (e.g., reproductive, maternal, newborn, and child health, infectious diseases, and noncommunicable diseases)

### 4 Supplementary Figures and Tables

#### 4.1 Supplementary Figures

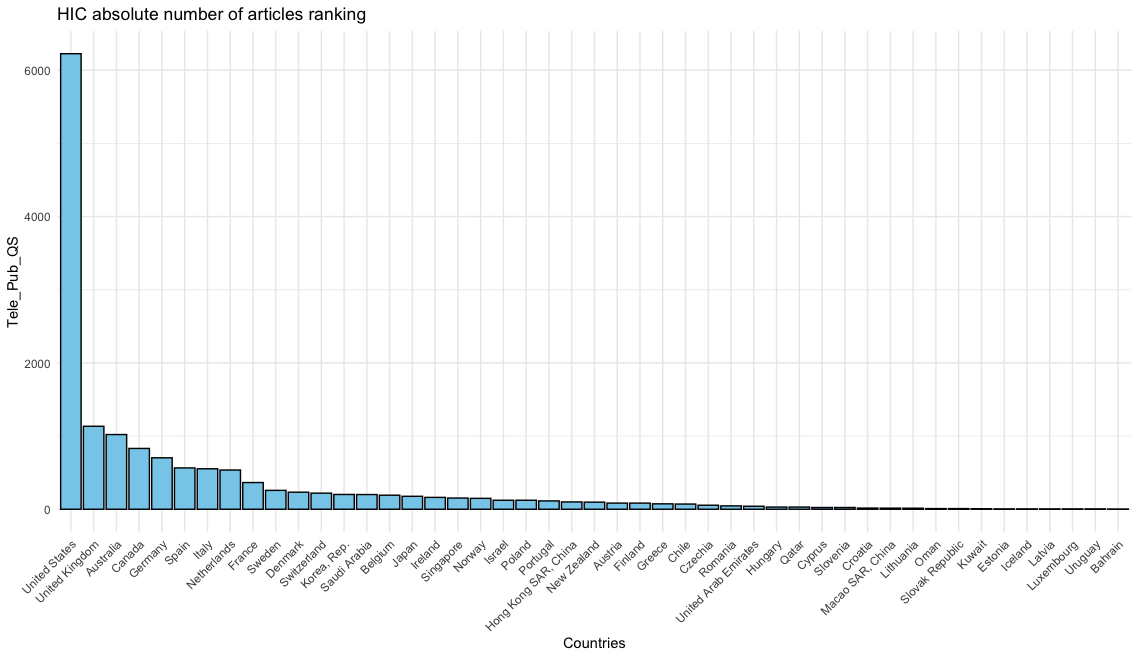

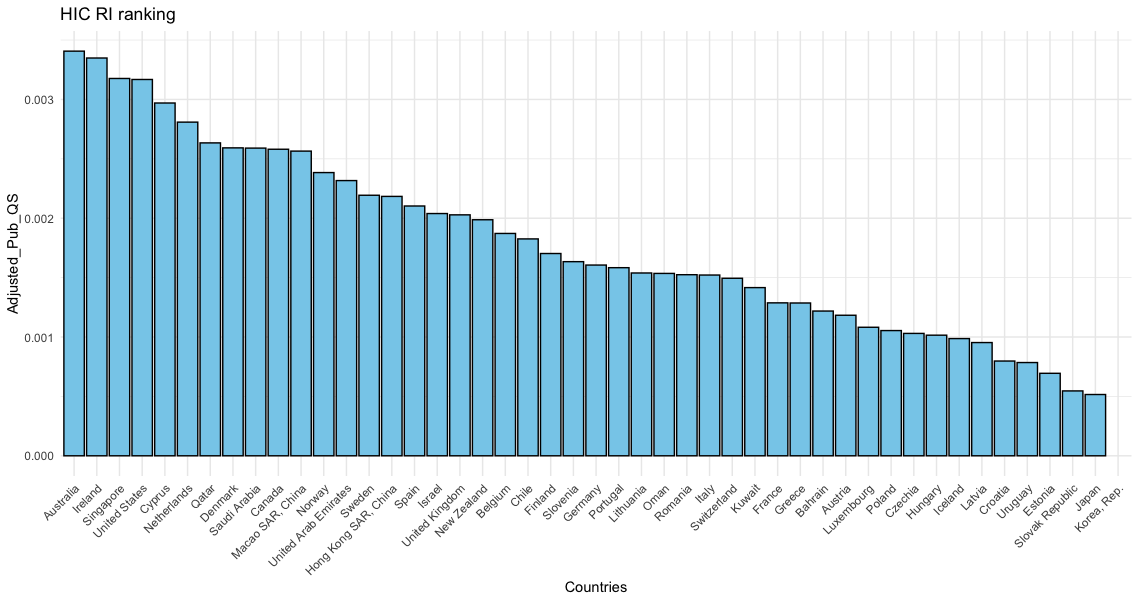

**Figure S1.** RI and absolute number of HIC telemedicine-related articles ranking

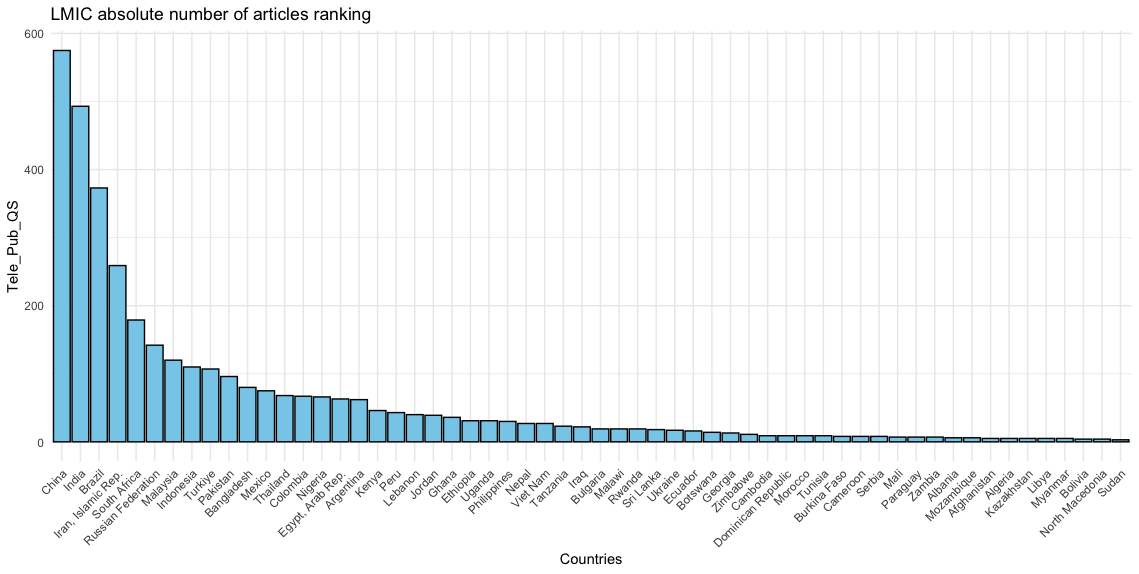

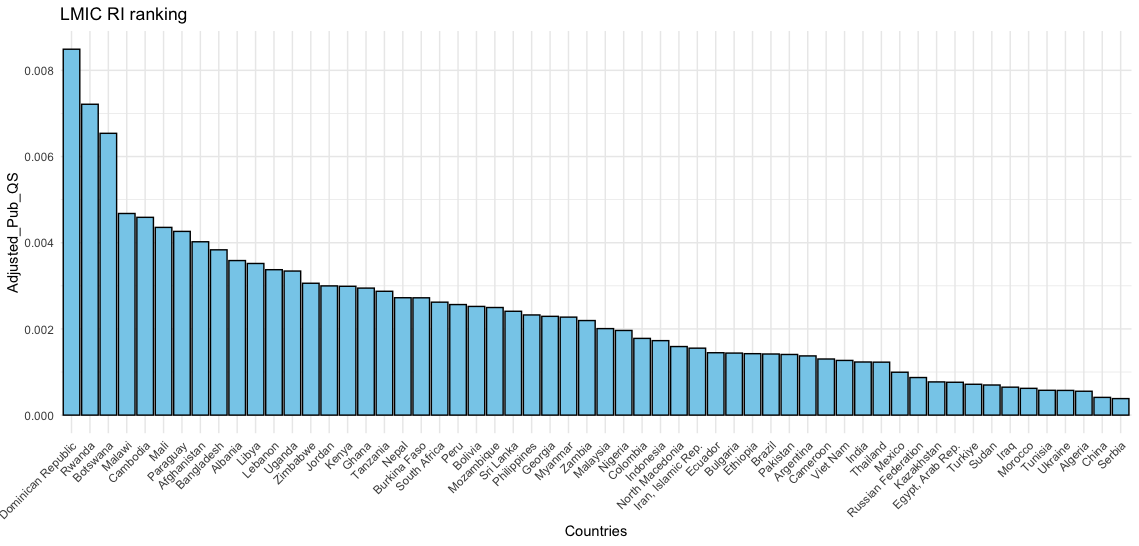

**Figure S2.** RI and absolute number of LMIC telemedicine-related articles ranking

#### 4.2 Supplementary Tables

**Table S1**. False negative screening, the number of telemedicine-related articles found in the Scopus profiles of the top 11 authors with the highest number of articles identified in our search

| Author's Name | Number of Articles found in search | Number of telemedicine related Articles found in their scopus profile |
| --- | --- | --- |
| Keely, Erin Joanne | 54 | 57 |
| Mehrotra, Ateev | 45 | 48 |
| Uscher-Pines, Lori | 31 | 35 |
| Liddy, Clare E. | 30 | 31 |
| Smith, Anthony C. | 30 | 36 |
| Mars, Maurice | 28 | 30 |
| Seto, Emily | 27 | 32 |
| Marcin, James Paul | 27 | 32 |
| Liddy, Clare | 27 | 29 |
| Caffery, Liam J. | 27 | 33 |
| Chavannes, N. Henrik | 26 | 30 |

**Table S2.** Descriptive comparison of bibliometric indices between HIC and LMIC, the pre- and post-COVID periods

| Outputs in top10% journal percentile | Outputs in top10% citation percentile | International collaboration | Citation per publication | FWCI^*^ | Scholarly output | Income and time classification |
| --- | --- | --- | --- | --- | --- | --- |
| 17% | 17% | 20.3% | 17 | 1.38 | 16581 | All |
| 16.5% | 20.1% | 22.9% | 26.5 | 1.38 | 3802 | All Pre Covid |
| 17.1% | 16.1% | 19.5% | 14.1 | 1.37 | 12,799 | All Post Covid |
| 13.4% | 16.6% | 43.0% | 15.8 | 1.2 | 4,242 | LMIC |
| 12.7% | 19.0% | 48.2% | 24.6 | 1.22 | 985 | LMIC Pre Covid |
| 13.6% | 15.9% | 41.2% | 13.1 | 1.19 | 3,257 | LMIC Post Covid |
| 18.5% | 18.1% | 23.6% | 18.1 | 1.46 | 13619 | HIC |
| 18.1% | 21.3% | 26.3% | 28.4 | 1.48 | 3,163 | HIC Pre Covid |
| 18.6% | 17.1% | 22.8% | 15 | 1.46 | 10,456 | HIC Post Covid |

^*^ Field-weighted citation impact

**Table S3.** Comparison of publication volume and FWCI across different timeframes and income groups

| **Classifications** | | **Tele Pub All** | | **Tele Pub QS** | | **Med Pub QS** | | **Adjusted Pub QS (RI)** | |
| --- | --- | --- | --- | --- | --- | --- | --- | --- | --- |
|  |  | **Cliff’s delta*** | **P-value** | **Cliff’s delta** | **P-value** | **Cliff’s delta** | **P-value** | **Cliff’s delta** | **P-value** |
| **All** | **Pre-COVID** |  |  |  |  |  |  | 0.5 (0.33-0.64) | <0.001 |
|  | **Post-COVID** |  |  |  |  |  |  |  |  |
| **HIC** | **Pre-COVID** |  |  |  |  |  |  | 0.65 (0.42-0.8) | <0.001 |
|  | **Post-COVID** |  |  |  |  |  |  |  |  |
| **LMIC** | **Pre-COVID** |  |  |  |  |  |  | 0.39 (0.13-0.6) | <0.001 |
|  | **Post-COVID** |  |  |  |  |  |  |  |  |
| **All** | **HIC** | 0.37 (0.14-0.56) | 0.001 | 0.38 (0.16-0.57) | 0.001 | 0.41 (0.2-0.59) | <0.001 | -0.14(-0.33 -0.06) | 0.21 |
|  | **LMIC** |  |  |  |  |  |  |  |  |
| **Pre-COVID** | **HIC** | 0.49 (0.24-0.69) | <0.001 | 0.49 (0.23-0.69) | <0.001 | 0.44 (0.18-0.65) | 0.001 | 0.06(-0.18-0.3) | 0.649 |
|  | **LMIC** |  |  |  |  |  |  |  |  |
| **Post-COVID** | **HIC** | 0.38 (0.14-0.57) | 0.001 | 0.41 (0.17-0.59) | 0.001 | 0.41 (0.19-0.59) | <0.001 | -0.1(-0.3-0.11) | 0.404 |
|  | **LMIC** |  |  |  |  |  |  |  |  |
| **Classifications** | | **Tele FWCI All** | | **Tele FWCI QS** | | **Med FWCI QS** | |  | |
|  |  | **Cliff’s delta** | **P-value** | **Cliff’s delta** | **P-value** | **Cliff’s delta** | **P-value** |  |  |
| **All** | **Pre-COVID** | 0.04(-0.15-0.23) | 0.647 | 0.09(-0.09-0.28) | 0.092 | -0.03(-0.22-0.16) | 0.981 |  |  |
|  | **Post-COVID** |  |  |  |  |  |  |  |  |
| **HIC** | **Pre-COVID** | -0.14(-0.38-0.11) | 0.626 | 0.09(-0.18-0.34) | 0.085 | 0.22(-0.11-0.55)^#^ | 0.189 |  |  |
|  | **Post-COVID** |  |  |  |  |  |  |  |  |
| **LMIC** | **Pre-COVID** | 0.21(-0.07-0.45) | 0.296 | 0.14(-0.13-0.39) | 0.432 | -0.05(-0.31-0.21) | 0.314 |  |  |
|  | **Post-COVID** |  |  |  |  |  |  |  |  |
| **All** | **HIC** | 0.28 (0.06-0.48) | 0.014 | 0.26 (0.05-0.45) | 0.023 | 0.5 (0.29-0.66) | <0.001 |  |  |
|  | **LMIC** |  |  |  |  |  |  |  |  |
| **Pre-COVID** | **HIC** | 0.48 (0.21-0.67 | <0.001 | 0.35 (0.07-0.57) | 0.011 | 0.49 (0.24-0.67) | <0.001 |  |  |
|  | **LMIC** |  |  |  |  |  |  |  |  |
| **Post-COVID** | **HIC** | 0.2(0-0.4) | 0.084 | 0.25 (0.03-0.45) | 0.033 | 0.49 (0.29-0.65) | <0.001 |  |  |
|  | **LMIC** |  |  |  |  |  |  |  |  |

*Cliff’s delta interpretation zones are as follows: cliff’s delta values below 0.15 were considered negligible (red), values between 0.15 and 0.32 indicated a small difference (orange), values between 0.33 and 0.46 represented a medium difference (yellow), and values greater than 0.46 reflected a large difference (green) (Meissel & Yao, 2024).

\# A paired t-test was employed for this comparison due to the normality of both the Med FWCI QS before and after COVID-19. Cohen’s d was reported as the measure of effect size for this analysis instead of Cliff’s Delta.

**Table S4.** Descriptive comparison of bibliometric indices across WHO regions and Iran

| Overall ranking | Quality ranking | Outputs in top10% journal percentile | Outputs in top10% citation percentile | International collaboration | Citations per publication | FWCI^*^ | Scholarly outputs | Geographical classification |
| --- | --- | --- | --- | --- | --- | --- | --- | --- |
| 1 | 2 | 19.8% | 18.1% | 21.1% | 18.9 | 1.56 | 7,942 | Americas with U.S. |
| 6 | 6 | 15.6% | 15.4% | 41.7% | 15.2 | 1.18 | 1,680 | Americas without U.S. |
| 3 | 4 | 16.3% | 16.6% | 35.2% | 17 | 1.3 | 5,463 | European |
| 4 | 3 | 13.5% | 22.0% | 51.9% | 20.5 | 1.5 | 997 | EMRO |
| 5 | 5 | 16.5% | 13.4% | 75.5% | 14.7 | 1.17 | 568 | African |
| 2 | 1 | 20.4% | 24.0% | 42.9% | 21.8 | 1.64 | 2,582 | Western Pacific |
| 7 | 7 | 10.4% | 14.2% | 36.6% | 14.5 | 1.18 | 1,072 | South-East Asia |
|  |  | 8.9% | 14.4% | 26.7% | 18.4 | 1.22 | 292 | Iran |
|  |  | 17% | 17% | 20.3% | 17 | 1.38 | 16581 | All |

^*^ Field-weighted citation impact

**Table S5.** Quality-based ranking of regions using TOPSIS across selected indices

| **Population** | **FWCI** | **citation_per_publication** | **ten_percentile_citation** | **ten_percentile_journal** | **Quality_score** | **Rank** |
| --- | --- | --- | --- | --- | --- | --- |
| **Western Pacific** | 1.64 | 21.8 | 0.24 | 0.204 | 1 | 1 |
| **Americas with USA** | 1.56 | 18.9 | 0.181 | 0.198 | 0.670 | 2 |
| **EMRO** | 1.5 | 20.5 | 0.22 | 0.135 | 0.585 | 3 |
| **European** | 1.3 | 17 | 0.166 | 0.163 | 0.424 | 4 |
| **African** | 1.17 | 14.7 | 0.134 | 0.165 | 0.319 | 5 |
| **Americas without USA** | 1.18 | 15.2 | 0.154 | 0.156 | 0.317 | 6 |
| **South-East Asia** | 1.18 | 14.5 | 0.142 | 0.104 | 0.043 | 7 |

**Table S6.** Overall ranking of regions using TOPSIS across selected indices

| **Population** | **quality** | **quantity** | **international_collab** | **Overall_score** | **rank** |
| --- | --- | --- | --- | --- | --- |
| **Americas with USA** | 0.67 | 7942 | 0.211 | 0.73 | 1 |
| **Western Pacific** | 1 | 2582 | 0.429 | 0.55 | 2 |
| **European** | 0.42 | 5463 | 0.352 | 0.53 | 3 |
| **EMRO** | 0.59 | 997 | 0.519 | 0.34 | 4 |
| **African** | 0.32 | 568 | 0.755 | 0.25 | 5 |
| **Americas without USA** | 0.32 | 1680 | 0.417 | 0.22 | 6 |
| **South-East Asia** | 0.04 | 1072 | 0.366 | 0.08 | 7 |

**Table S7.** Results of correlation tests between socioeconomic indicators and telemedicine-related output, using both absolute number of publications and research interest (RI)

| **Socioeconomic Indicators** | **Category** | **Absolute Number** | | **Research Interest** | | **Absolute number^$^** | **Research Interest** |
| --- | --- | --- | --- | --- | --- | --- | --- |
|  |  | **r** | **P-value** | **r** | **P-value** |  |  |
| Health Expenditure (percentage of GDP) | LMIC | -0.2 | 0.13 | 0.06 | 0.63 |  |  |
|  | HIC | 0.66 | <0.001 | 0.16 | 0.31 | + |  |
| Health Expenditure per Capita (PPP) | LMIC | 0.24 | 0.07 | -0.32^#^ | 0.01 |  | - |
|  | HIC | 0.63 | <0.001 | 0.45^#^ | 0.002 | + | + |
| HDI | LMIC | 0.29 | 0.03 | -0.38 | 0.003 | + | - |
|  | HIC | 0.5 | <0.001 | 0.47 | 0.001 | + | + |
| R&D expenditure (percentage of GDP) | LMIC | 0.55 | <0.001 | -0.40 | 0.02 | + | - |
|  | HIC | 0.56 | <0.001 | 0.1 | 0.51 | + |  |
| R&D Researchers | LMIC | 0.28 | 0.14 | -0.59^#^ | <0.001 |  | - |
|  | HIC | 0.44 | 0.003 | 0.15^#^ | 0.33 | + |  |
| Electricity accessibility (percentage of Rural population) | LMIC | 0.22 | 0.1 | -0.48 | <0.001 |  | - |
|  | HIC | NA | NA | NA | NA |  |  |
| Hospital Beds^*^ | LMIC | 0.14 | 0.45 | -0.24 | 0.17 |  |  |
|  | HIC | 0.02 | 0.9 | -0.49 | <0.001 |  | - |
| Population Using Internet (percentage of total population) | LMIC | 0.23 | 0.08 | -0.33^#^ | 0.01 |  | - |
|  | HIC | 0.1 | 0.52 | 0.36^#^ | 0.01 |  | + |
| Secure Internet Server | LMIC | 0.35 | 0.007 | -0.30 | 0.02 | + | - |
|  | HIC | 0.31 | 0.04 | 0.16 | 0.29 | + |  |
| Secure Internet Server All | LMIC | 0.65 | <0.001 | -0.47 | <0.001 | + | - |
|  | HIC | 0.8 | <0.001 | 0.2 | 0.1922 | + |  |
| Land Area | LMIC | 0.32 | 0.01 | -0.42 | 0.001 | + | - |
|  | HIC | 0.54 | <0.001 | 0.03 | 0.82 | + |  |
| Population Density^*^ | LMIC | 0.35 | 0.007 | 0.09 | 0.5 | + |  |
|  | HIC | 0.05 | 0.73 | 0.04 | 0.8 |  |  |
| High Technology Exports | LMIC | 0.37 | 0.005 | -0.45 | <0.001 | + | - |
|  | HIC | 0.43 | 0.003 | -0.08 | 0.02 | + |  |
| Mobile Cellular Subscription | LMIC | 0.15 | 0.26 | -0.16 | 0.23 |  |  |
|  | HIC | -0.35 | 0.015 | -0.07 | 0.66 | - |  |
| Nurses and Midwife^*^ | LMIC | 0.40 | 0.004 | -0.26 | 0.08 | + |  |
|  | HIC | 0.37 | 0.02 | 0.17 | 0.3 | + |  |
| Physicians^*^ | LMIC | 0.36 | 0.01 | -0.4^#^ | 0.005 | + | - |
|  | HIC | 0.02 | 0.9071 | -0.21^#^ | 0.18 |  |  |
| Population aged 65 and above (percentage of total population) | LMIC | 0.25 | 0.06 | -0.45 | <0.001 |  | - |
|  | HIC | 0.23 | 0.1213 | -0.38 | 0.009 |  | - |
| Rural Population (percentage of total population) | LMIC | -0.28 | 0.03 | 0.24 | 0.07 | - |  |
|  | HIC | -0.01 | 0.93 | -0.3 | 0.048 |  | - |
| Population in Largest cities (percentage of urban population) ^*^ | LMIC | -0.42 | 0.001 | 0.45 | <0.001 | - | + |
|  | HIC | -0.6 | <0.001 | -0.08 | 0.6 | - |  |
| UHC Service Index | LMIC | 0.35 | 0.007 | -0.45^#^ | <0.001 | + | - |
|  | HIC | 0.58 | <0.001 | 0.21^#^ | 0.16 | + |  |
| Literacy Rate | LMIC | 0.17 | 0.23 | -0.29^#^ | 0.04 |  | - |
|  | HIC | -0.16 | 0.49 | -0.39^#^ | 0.08 |  |  |
| ICT Export (percentage of total goods) | LMIC | 0.42 | 0.002 | -0.33 | 0.01 | + | - |
|  | HIC | 0.13 | 0.38 | 0.003 | 0.98 |  |  |
| ICT Import (percentage of total goods) | LMIC | 0.55 | <0.001 | -0.37 | 0.006 | + | - |
|  | HIC | 0.25 | 0.1 | 0.21 | 0.16 |  |  |
| ICT service Export (percentage of service exports) | LMIC | 0.14 | 0.31 | -0.12 | 0.4 |  |  |
|  | HIC | -0.1 | 0.51 | -0.14 | 0.35 |  |  |
| ICT service Export (in US dollar) | LMIC | 0.63 | <0.001 | -0.55 | <0.001 | + | - |
|  | HIC | 0.72 | <0.001 | 0.33 | 0.03 | + | + |

*These indicators are expected to be negatively correlated with the absolute number of publications and RI.

#Unlike other indicators, the correlation coefficient(r) here is derived from the Pearson correlation test because of the normal distribution of RI and the indicator

$The strength of the relationships is represented by the following colors, with the direction indicated by “+” or “-“: red for negligible strength/ orange for weak to low strength/ yellow for moderate to low strength/ light green for moderate strength/ green for strong strength/ dark green for very strong strength/ shading for no relationship.

**Table S8.** Classification of indicators across HIC and LMIC based on their relationship with the absolute number of articles, adjusted number of articles (RI), both, or neither, and their directionality

| **LMIC** | | | | | **HIC** | | | | |
| --- | --- | --- | --- | --- | --- | --- | --- | --- | --- |
| **Group A** | | **Group C** | **Group D** | **Group E** | **Group B** | | **Group C** | **Group D** | **Group E** |
| **Both Absolute number and research interest** | | **Only absolute number^#^** | **Only research interest** | **none** | **Both Absolute number and research interest** | | **Only absolute number** | **Only research interest** | **none** |
| **Same direction*** | **Opposite direction*** |  |  |  | **Same direction** | **Opposite direction** |  |  |  |
|  | HDI | Population Density | Health Expenditure per Capita (PPP) | Health Expenditure (percentage of GDP) | Health Expenditure per Capita (PPP) |  | Health Expenditure (percentage of GDP) | Hospital Beds | Electricity accessibility (percentage of Rural population) |
|  | R&D expenditure (percentage of GDP) | Nurses and Midwife | R&D Researchers | Hospital Beds | HDI |  | R&D expenditure (percentage of GDP) | Population Using Internet (percentage of total population) | Population Density |
|  | Secure Internet Server | Rural Population (percentage of total population) | Electricity accessibility (percentage of Rural population) | Mobile Cellular Subscription | ICT service Export (in US dollar) |  | R&D Researchers | Population aged 65 and above (percentage of total population) | Physicians |
|  | Secure Internet Server All |  | Population Using Internet (percentage of total population) | ICT service Export (percentage of service exports) |  |  | Secure Internet Server | Rural Population (percentage of total population) | Literacy Rate |
|  | Land Area |  | Population aged 65 and above (percentage of total population) |  |  |  | Secure Internet Server All |  | ICT Export (percentage of total goods) |
|  | High Technology Export |  | Literacy Rate |  |  |  | Land Area |  | ICT Import (percentage of total goods) |
|  | Physicians |  |  |  |  |  | High Technology Export |  | ICT service Export (percentage of service exports) |
|  | Population in Largest cities (percentage of urban population) |  |  |  |  |  | Mobile Cellular Subscription |  |  |
|  | UHC service index |  |  |  |  |  | Nurses and Midwife |  |  |
|  | ICT Export (percentage of total goods) |  |  |  |  |  | Population in Largest cities (percentage of urban population) |  |  |
|  | ICT Import (percentage of total goods) |  |  |  |  |  | UHC service index |  |  |
|  | ICT service Export (in US dollar) |  |  |  |  |  |  |  |  |

* Same direction: The correlation coefficient of the absolute number of publications with the indicator and the correlation coefficient of the RI with the same indicator are both positive or both negative.

* Opposite direction: One of the correlation coefficients of the absolute number of publications and RI with a same indicator is negative and the other is positive.

#Indicators highlighted in light yellow are correlated in the opposite direction to expectations, while those highlighted in light green are correlated in the expected direction

**Table S9.** Studies investigating socioeconomic drivers in other research fields than telemedicine

| **Study** | **Subject** | **Socioeconomic driver** | **Bibliometric indicator** |
| --- | --- | --- | --- |
| (C. Wen et al., 2024) | Total anomalous pulmonary venous connection | - GDP (R = 0.887) - Research & development (R&D) expenditure (R = 0.375) - Population (R = 0.694) - Journals (R = 0.751) | National publications |
|  |  | - GDP (R = 0.881) - R&D expenditure (R = 0.446) - Population (R = 0.305) - Journals (R = 0.917) | National citations |
| (Ali et al., 2024) | spontaneous perineal tears sustained during [childbirth](https://www.sciencedirect.com/topics/medicine-and-dentistry/childbirth) | - GDP per capita(positive) - Average Health Expenditure(positive) | Number of articles |
| (Zhang et al., 2024) | intracranial aneurysm management with artificial intelligence technology | - GDP(positive) | - Number of articles - Number of citations |
| (Zheng et al., 2024) | orthodontic tooth movement | - global GDP(r = 0.915) - GDP of individual countries/regions(r = 0.976) | Number of articles |
| (Koleen C. Pasamba & Jean Anne B. Toral, 2024) | preterm birth in Southeast Asia | - GDP per capita allocated to R&D)positive) | Number of articles |
| (Li, 2023) | educational research from Singapore, Japan, and South Korea | - R&D expenditures(Positive) - R&D expenditures as a percentage of GDP(Positive) | Annual number of articles in South Korea |
| (Dündar et al., 2023) | acetabular fractures | - GDP(R = 0.719) - GDP per capita(R = 0.701) | Number of articles |
| (Grillo et al., 2023) | Middle Eastern oral and maxillofacial surgery | - Total population (R = 0.6052) - Land area (R = 0.302291) - HDI(R = 0.1747) | Number of articles |
| (Bitar et al., 2024) | Congenital heart disease in the Arab world | - GDP(positive) | Number of articles |
| (Ahn et al., 2023) | Myopia research in East Asia | - GDP(positive) | Annual number of articles for only China and South Korea |
| (Sater et al., 2023) | Cancer research in MENA | No correlation with GDP per capita | Number of articles |
| (Awada et al., 2023) | Bladder cancer research | - GDP(positive) | Number of articles |
| (Apor & Jamora, 2022) | the research output of adult and child neurologists in the Philippines | - Population(positive) - GDP(positive) - Health expenditure(positive) - Number of healthcare establishments(positive) - Number of neurologists(positive) - Number of research personnel(positive) | Number of articles |
| (Omar et al., 2022) | Neurosurgical research in Southeast Asia | - percentage of the GDP allocated to R&D (positive) - Number of collaborations (positive) | - number of publications - total citations - H-index - i(10)-index |
| (Sánchez-Marqués et al., 2022) | Schistosomiasis research during COVID-19 | - Population(Positive) - Gross domestic expenditure on R&D(GERD)(Positive) - Researchers per million inhabitants(positive) - GDP per capita (no relation) | Number of articles |
| (X. Yang et al., 2022) | Cathepsin B research | - GDP(R=0.9745) | Number of articles |
| (H.-Y. Yang et al., 2022) | ERCP research | - GDP(R=0.870) | Number of articles |
| (P. Wen et al., 2022) | Gout | - GDP(positive) - International collaboration(positive) | Number of articles |
| (Chanbour et al., 2021) | Neurosurgery research in Arab countries | - GDP(not related) - Population(R squared = 0.49) | Number of articles |
| (Lerman et al., 2021) | OECD internal medicine research | - Health expenditure percentage of GDP(H-index, r = 0.75; number of citations, r = 0.72; number of articles, r = 0.62; and number of citable articles, r = 0.61) - GERD(number of citations, r = 0.6; H-index, r = 0.6; number of articles, r = 0.53; number of citable articles, r = 0.51) - GDP per capita(number of citations, r = 0.46; citations per article, r = 0.54; H-index, r = 0.5) | - H-index - Number of citations - Number of articles - Number of citable articles - Number of citations per article |
| (Koelmel et al., 2015) | Phytotechnologies for remediation | - HDI(negative) | Proportion of phytotechnologies research(research interest) |
| (Lerman et al., 2021) | Ophthalmology research in OECD | - Health expenditure(H-index, *r* = 0.711; number of articles, *r* = 0.589; number of citable articles, *r* = 0.593; number of citations, *r* = 0.673) - GERD(H-index, *r* = 0.564)   Regression analysis also conducted and confirmed the correlations | - H-index - Number of articles - Number of citable articles - Number of citations |
